# Public perceptions and experiences of social distancing and social isolation during the COVID-19 pandemic: A UK-based focus group study

**DOI:** 10.1101/2020.04.10.20061267

**Authors:** Simon N Williams, Christopher J. Armitage, Tova Tampe, Kimberly Dienes

## Abstract

**OBJECTIVE:** Explore the perceptions and experiences of the UK public of social distancing and social isolation measures related to the COVID-19 pandemic.

**DESIGN:** Qualitative study comprising five focus groups carried out online during the early stages of the UK’s social distancing and isolation measures (5-12 days post lockdown).

**SETTING:** Online video-conferencing

**PARTICIPANTS:** 27 participants, all UK residents aged 18 years and older, representing a range of gender, ethnic, age and occupational backgrounds.

**RESULTS:** The social distancing and isolation associated with COVID-19 policy has had having substantial negative impacts on the mental health and wellbeing of the UK public within a short time of policy implementation. It has disproportionately negatively affected those in low-paid or precarious employment. Practical social and economic losses - the loss of (in-person) social interaction, loss of income and loss of structure and routine – led to psychological and emotional ‘losses’ – the loss of motivation, loss of meaning, and loss of self-worth. Participants reported high adherence to distancing and isolation guidelines but reported seeing or hearing of non-adherence in others. A central concern for participants was the uncertainty duration of the measures, and their ability to cope longer-term. Some participants felt they would have lingering concerns over social contact while others were eager to return to high levels of social activity.

**CONCLUSIONS:** A rapid response is necessary in terms of public health programming to mitigate the mental health impacts of COVID-19 social distancing and isolation. Initial high levels of support for, and adherence to, social distancing and isolation is likely to wane over time, particularly where end dates are uncertain. Social distancing and isolation ‘exit strategies’ must account for the fact that, although some individuals will voluntarily or habitually continue to socially distance, others will seek high levels of social engagement as soon as possible.

**What is already known on this topic**

- Adherence to non-pharmaceutical interventions during pandemics is lower where people have low trust in government and where people perceive themselves at relatively low risk from the disease
- There is a need for evidence on public perceptions and experiences of the psychological and social public experiences of COVID-19 related social distancing and isolation, and its relation to adherence.

**What this study adds**

- People lack trust in government and perceive themselves at low personal risk,but closely adhere to social distancing and isolation measures motivated by social conscience, and are critical of non-adherence in others.
- Population-wide social distancing and isolation can have significant negative social and psychological impacts within a short time of policy implementation.
- Key concerns during social distancing and isolation are uncertainty of duration and ability to cope longer-term.
- At the end of pandemic ‘lockdowns’, some individuals will likely voluntarily or habitually continue to socially distance, while others will likely seek high levels of social engagement as soon as possible.

## INTRODUCTION

The current coronavirus (COVID-19) pandemic presents the greatest threat to public health in living memory and promises to be the deadliest pandemic since 1918-19.[1-2] Pandemics are challenging for clinical and public health agencies and policymakers because of the scientific and medical uncertainty that accompanies novel viruses like COVID-19.[3-4] Since COVID-19 is a new virus, pharmaceutical interventions like vaccines are not presently available. Public health policy is therefore exclusively reliant on non-pharmaceutical interventions (NPIs). The key NPIs being used in relation to the COVID-19 pandemic in the UK and globally (in addition to personal hygiene advisories (e.g. emphasising regular and thorough handwashing)) are *social distancing* (e.g. prohibiting public gatherings, closing schools and other non-essential services, and keeping a distance of >2 metres apart from others) and *social isolation* (e.g. remaining indoors except for one brief outing for per day for physical activity or ‘essential supplies’).[5]

Due to the unprecedented scale and severity of the social distancing and social isolation measures being implemented in response to COVID-19, the social and psychological impacts on the public are also likely to be unprecedented in scale and severity. A recent rapid review of the psychological impact of quarantine found that longer quarantine duration, infection fears, frustration and boredom, inadequate supplies, inadequate information, financial loss and stigma were among the major stressors.[6] Another systematic review of the literature on NPIs in relation to pandemic influenza and SARS found that people actively evaluate NPIs in terms of criteria such as perceived necessity, efficacy, acceptability and feasibility.[7] Public views on social distancing and social isolation are ambivalent in some contexts because of their perceived adverse social and economic impacts and their ability to attract stigma, particularly amongst those required to self-isolate.[7] Existing research on social distancing and isolation highlights a number of challenges for public health policymakers, including a lack of trust in government;[8] concerns over strains in family resources;[8] gaps and confusions in some areas of pandemic information communication;[9] and low adherence to voluntary social isolation and relatively low adherence to non-attendance at public gatherings.[10] Although there is existing research from past pandemics on its likely effects,[6] and new quantitative research is starting to emerge,[11] there is no published qualitative evidence on public perceptions and experiences of the psychological and social public experiences of COVID-19 related social distancing and social isolation, and its relation to adherence – a gap that the present study addresses.

This study aimed to explore four main questions: (1) What are the social and psychological impacts of social distancing and isolation experienced by the UK public during the COVID-19 pandemic? (2) What are people’s views on government communication around social distancing and isolation? (3) What are people’s current experiences of adherence in relation to social distancing and isolation? (4) What are people’s views on the future in regard to COVID-19 social distancing and isolation. This study therefore aims to contribute to knowledge of adherence to social distancing and isolation policy to provide insight into how communication with the public on social distancing and isolation may be shaped and improved in the future.

### PARTICIPANTS AND METHODS

Five online focus groups with 27 participants were run between March 28^th^ and April 4^th^, 2020 (5-12 days after lockdown commenced on the 23^rd^ March 2020). Participants were adults aged 18 years or over currently residing in the UK. Under normal circumstances, online focus groups can be a useful way of eliciting public views related to matters of health and medicine, particularly from diverse and geographically dispersed participants [12-13] but were necessary due to social isolation policy.

Purposive sampling was used to provide a diverse range of ages, genders, race/ethnicities and social backgrounds and to explore the study’s research questions.[14-15] Potential participants were asked to complete a very brief demographic survey to provide background information and to facilitate recruitment of a diverse population (Table 1). Due to social distancing measures, it was necessary for all recruitment to be conducted online. Researchers used a combination of social media snowball sampling, online community and volunteer advertising sites and social media advertisements (Facebook ads).

**Table 1:**
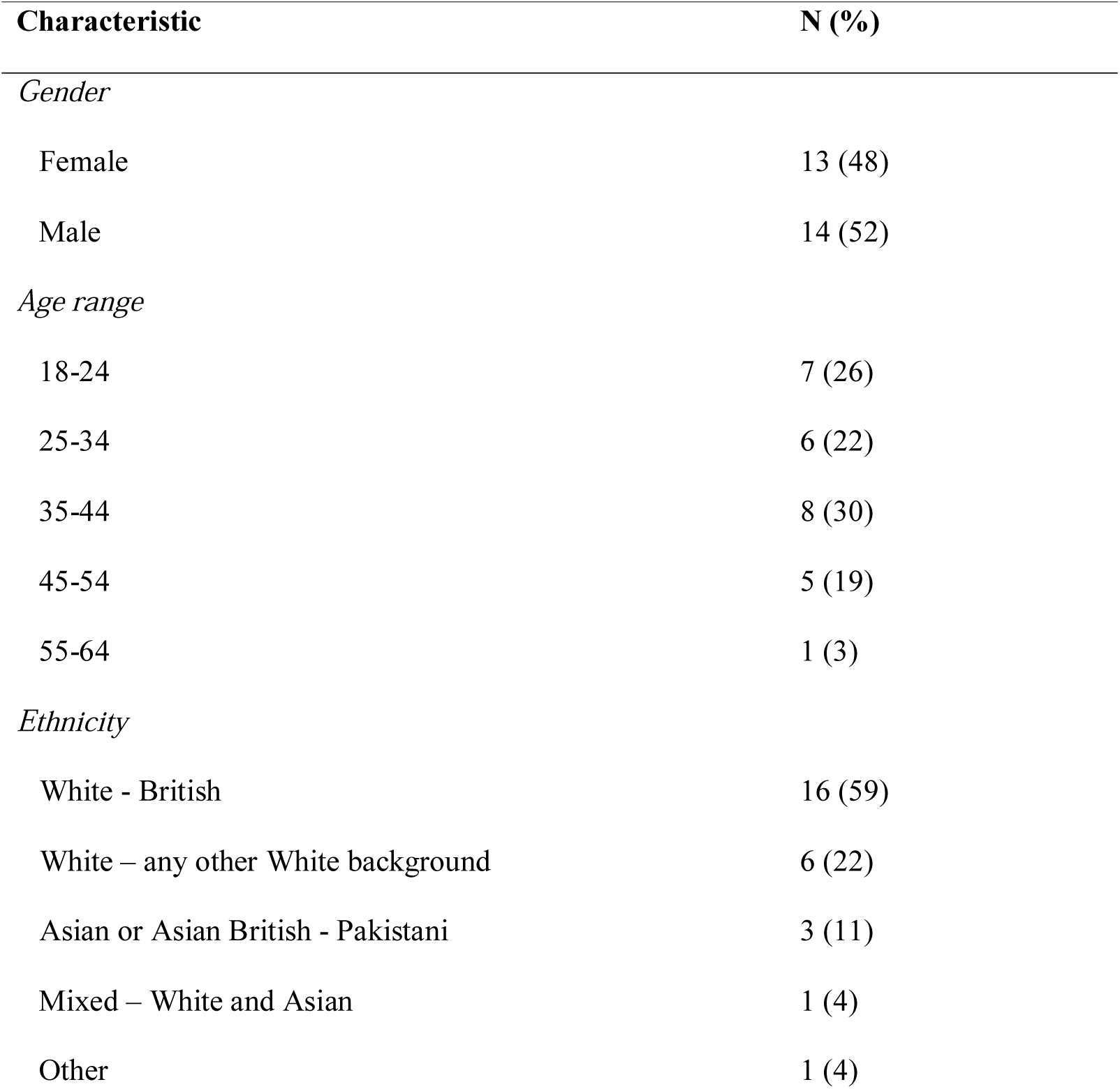
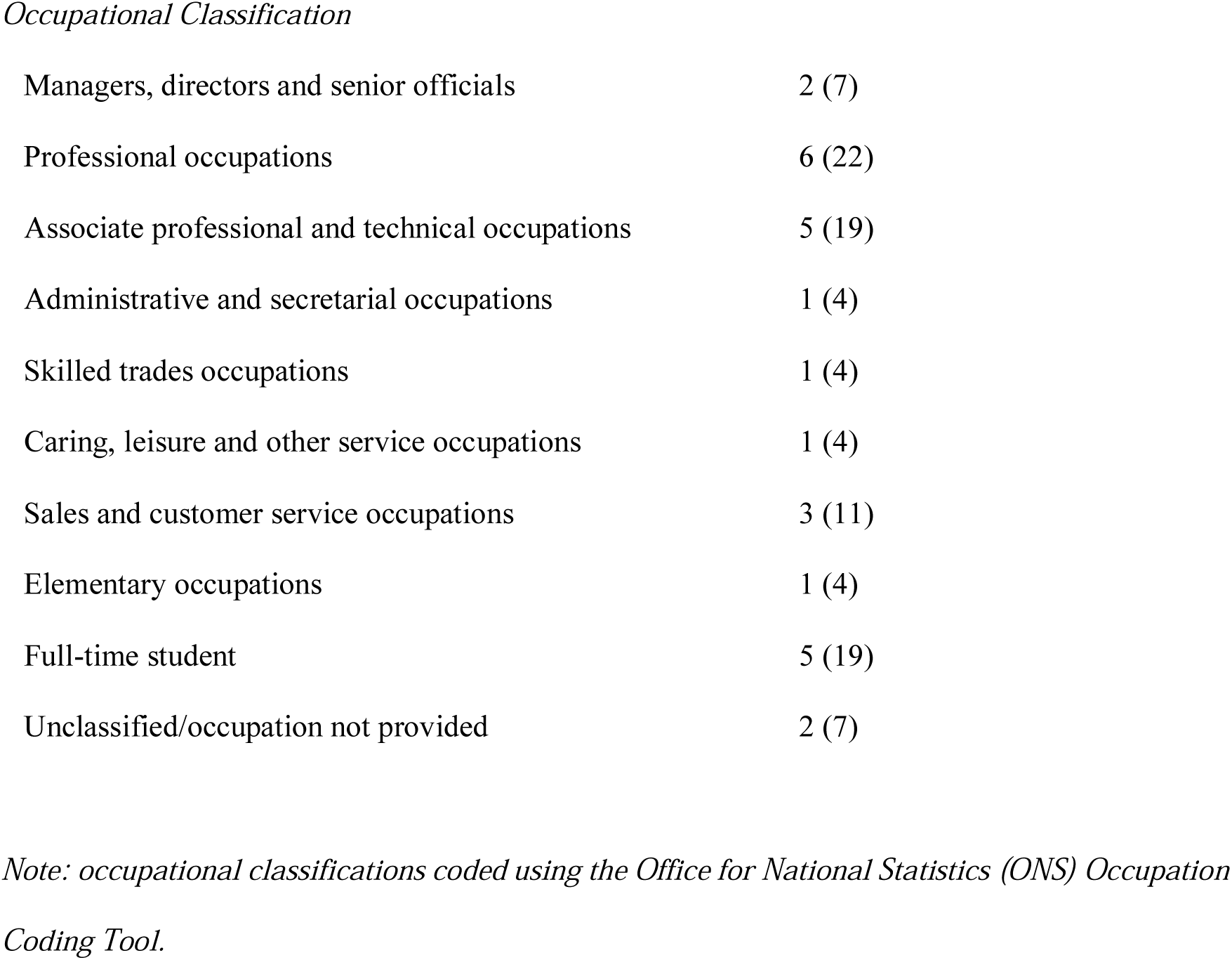
Demographic details reported by participants.

| Characteristic | N (%) |
| --- | --- |
| *Gender* |  |
| Female | 13 (48) |
| Male | 14 (52) |
| *Age range* |  |
| 18-24 | 7 (26) |
| 25-34 | 6 (22) |
| 35-44 | 8 (30) |
| 45-54 | 5 (19) |
| 55-64 | 1 (3) |
| *Ethnicity* |  |
| White - British | 16 (59) |
| White – any other White background | 6 (22) |
| Asian or Asian British - Pakistani | 3 (11) |
| Mixed – White and Asian | 1 (4) |
| Other | 1 (4) |
| *Occupational Classification* |  |
| Managers, directors and senior officials | 2 (7) |
| Professional occupations | 6 (22) |
| Associate professional and technical occupations | 5 (19) |
| Administrative and secretarial occupations | 1 (4) |
| Skilled trades occupations | 1 (4) |
| Caring, leisure and other service occupations | 1 (4) |
| Sales and customer service occupations | 3 (11) |
| Elementary occupations | 1 (4) |
| Full-time student | 5 (19) |
| Unclassified/occupation not provided | 2 (7) |

*Note: occupational classifications coded using the Office for National Statistics (ONS) Occupation Coding Tool.*

To ensure that online discussion was manageable, focus groups were kept to between 5-8 participants. Each group met virtually via a web video-conferencing platform (Zoom) for between 60-90 minutes. Participants joined using both video and audio. All focus groups were organised and moderated by SW (a medical social scientist). The topic guide for the focus groups was initially developed using existing literature on public attitudes and experiences in past pandemics and was tested and refined in a pilot focus group. The main topics for the focus groups were: general views on social distancing and isolation; health impacts of social distancing and isolation; views on government COVID-19 advice and communication; and views on compliance with, and the future impacts of, social distancing and isolation.

### Analysis

Data collection and analysis followed an iterative process, whereby emergent themes from early focus groups were used to add to or refine questions and prompts during subsequent focus groups. All focus groups were recorded and transcribed for coding. SW and KD analysed the transcripts and developed and applied the thematic coding framework. Themes were discussed and developed with CJA and TT during virtual research group meetings. To help analysis we looked to validate “sensitive moments” between groups that indicated difficult but important issues.[16] Negative case analysis was used to seek for information that did not fit emergent themes, and where this occurred, themes were modified accordingly.[17]. Following a grounded theory approach, data were organised into primary and more focused codes that provided insight into identified themes.[14,18] Data collection and analysis continued until saturation occurred (that is, until no new significant themes emerged).[18] Data were analysed in NVivo (version 11.4.3, QRS).

## RESULTS

Analysis revealed four broad themes: (1) the negative social and psychological impacts of social distancing and isolation during the COVID-19; (2) criticisms of government communication around social distancing and isolation; (3) current adherence and non-adherence of self and others; and (4) Uncertainty, social-reintegration and the future. Within each broad theme were a number of substantive sub-themes that are discussed below, supplemented by indicative quotes in Boxes 1-4.

### The social and psychological impacts of COVID-19 social distancing and isolation

All participants felt that the social distancing and isolation polices had had significant social and psychological impacts on their lives and the central theme was loss (Box 1). This experience of loss, which one participant likened to a process of “grieving” (Participant 25, male, age 58, London), consisted of three practical social and economic losses: loss of (in-person) social interaction, loss of income, and loss of structure and routine. These in turn led to three psychological and emotional “losses”: loss of motivation, loss of meaning, and loss of self-worth.

First, participants spoke of a loss of social interaction. The suddenness and extensiveness of the lack of face-to-face contact had, even after only one week of lockdown, already “taken its toll on mental health” (Participant 2, Male, age 31, Manchester), leaving participants feeling “alienated” (Participant 6, male, age 20, London). A number talked about feeling depressed or anxious as a result of social distancing or isolation, an experience some likened to “a prison” (Participant 8, female, age 40, London). Second, a number of participants discussed how a loss of income, either through permanent loss of a job, or through temporary loss (via lost clients or customers or being furloughed), had left them feeling “quite depressed” (Participant 8, female, age 40, London). Third, participants expressed of a loss of structure and routine. The inability to go to work, or for some the significant re-structuring of work patterns, including balancing home working with home schooling, meant that participants felt “overwhelmed” (Participant 9, female, age 34, London).

Participants discussed how impacts like losing their job or not being able to go to work, and not being able to socialise with friends, meant they experienced a general loss of meaning in life. One participant already felt in need of professional mental health support, less than two weeks into isolation. Participants also spoke of a loss of motivation to perform basic everyday tasks, such as personal hygiene and grooming or exercising. For some, this lack of motivation had left them feeling “sluggish” (Participant 5, male, age 26, London). Finally, participants expressed feeling a loss of self-worth. These emotional and psychological losses were particularly acute for those living in more urban, densely populated cities like London or Birmingham. They were also especially evident amongst those in low-paid or precarious occupations, who had either lost their job or income or were now relying on parental, familial or state financial support as a result of the pandemic.

#### Box 1

**The social and psychological impacts of COVID-19 social distancing and isolation**

*Loss of social interaction*

“I’ve been working at home for the past week and a bit and it’s taken its toll … because you think social contact is such an important part of everyday life and now it’s like you walk down the street and people are almost too scared to walk too close. It’s so alien.” (Participant 6, male, age 20, London)

“It’s all over the news, it’s all over your phone, it’s all over the TV, it’s basically everywhere you turn you are hearing about it. All of a sudden, we can’t do these things we used to do, like going to the shops and restaurants, and we just have to stay in, and I think people feel claustrophobic in both a physical and an emotional sense.” (Participant 5, male, age 26, London)

*Loss of structure and routine*

“I feel really lazy at home. I feel sluggish. I feel out of my routine. I feel much less active, both mentally and physically. You know, not taking the trip to work every day. My working from home schedule is neither here nor there. Mentally I am not as sharp, I feel like I am taking lots of naps in the day.” (Participant 5, male, age 26, London)

“I’m literally planning day-to-day as things go along. … I’m not used to having the kids every single day because they are usually at school. It’s difficult to work around them, I can’t do anything with them, because I can’t go out. I feel so scared and don’t want to risk it.” (Participant 9, Female, age 34, London)

*Loss of meaning*

“All this talk about social distancing and things is so depressing, terrible, I mean I have even been contemplating on contacting *The Samaritans* just to be able to try to get through all this.” (Participant 10, male, age 44, London)

“Being locked in a room trying to find something meaningful to do during the day, and I think it’s had a severe impact … I hope something changes within a few weeks, so I am able to go out and live a fulfilling life” (Participant 1, male, age 30, Birmingham)

*Loss of motivation*

“Physically it has had a toll on people. All day you are stuck at home. You eat, you sleep, you work, its gonna have an effect on the body, there is no real drive or motivation.” (Participant 6, male, age 20, London)

“We are feeling very down and demotivated, very low very depressed to some extent… it’s become more stressful to get by and function on a daily basis.” (Participant 1, male, age 30, Birmingham)

*Loss of self-worth*

“Your self-worth goes down a bit, because you can’t socialise with people and make yourself feel good about yourself.” (Participant 2, male, age 31, Manchester)

“The company I work for has closed down and I have had to apply for welfare assistance … and I’ve had to go and live with my parents now, and they have had to support me financially. … it’s been difficult, the whole mental health, the ability to function and get by, and being constantly locked in.” (Participant 1, male, age 30, Birmingham)

### Criticisms of government communication around social distancing and isolation

Most participants felt that guidance on social distancing and isolation had been generally unclear, although some described how it had “become clearer” over the course of the pandemic (Participant 7, male, age 20, Glasgow) (Box 2). Many participants exhibited a lack of trust either in government, who were seen to be “politicising” the pandemic (Participant 22, male, age 51, North-West England), or in the media, who they felt were providing confusing information or “mixed messages” (Participant 1, male, age 30, Birmingham). Participants felt that despite being locked at home, the constant media and social media attention on COVID-19 made them feel “claustrophobic in both a physical and an emotional sense” (Participant 5, male, age 26, London), and that “seeing others in a heightened state of anxiety makes it harder to suppress that in yourself” (Participant 21, female, age 46, North West England).

Another common criticism was over the ambiguity of terms such as ‘essential’ and ‘emergency’ supplies and services. This ambiguity, participants argued, meant that advice was either hard to follow or implement, or that “loopholes” could be exploited (Participant 19, female, age 21, South Wales) (see section below on compliance, non-compliance and the future).

#### Box 2

**Views on government communication around social distancing and isolation**

*Mixed or unclear messages*

“After reading several news publications and channels, there has been much campaign around social distancing, and with isolation you normally associate it when you have got the virus yourself, but I think over the past week there have been several mixed messages over social distancing.” (Participant 1, male, age 30, Birmingham)

“I’m trying to pick my way through what is happening, a lot of politicians are politicising it [COVID-19] and when you read the internet, it is very difficult to know what is real, true or valid, even if you read a broad church of views, facts and figures, it is still very difficult to make sense of it all (Participant 22, male, age 51, North-West England).

*Ambiguous definitions*

“Now everyone has been told that they have to stay in their houses, and people are thinking well ‘this can be classed as essential, and this can be classed as essential, whereas although we have been told a list of things we can do, people are finding loopholes and finding ways to get round them” (Participant 19, female, age 21, South Wales)

“I have seen loads of people outside, and I wonder how people will enforce that [penalty fines for not social distancing], I’m wondering how can someone prove they are going for an ‘emergency reason’?” (Participant 2, male, age 31, Manchester)

### Current adherence and non-adherence of self and others

All participants reported being highly adherent to government instructions on social distancing (Box 3). Participants described how, despite the perceived lack of clarity discussed above, they had been social distancing and isolating as far as possible. Participants also displayed a high degree of social consciousness, with many acknowledging that despite not perceiving themselves as being at high risk, they were doing it to “save lives” and protect those most vulnerable to the disease.

Despite reporting their own high degree of adherence, many participants suggested that they had seen instances of non-adherence in others. Observations of non-adherence were associated with three main factors. First, non-adherence was seen to be due to a lack of social conscience. Participants were generally critical of what they perceived to be a minority of “inconsiderate” (Participant 27, female, age 46, London) or “arrogant” (Participant 17, male, age 22, South Wales) individuals who were not observing instructions related, for example, to public gatherings and not keeping a distance of >2 metres apart from others when out for daily walks or runs. Second, non-adherence was seen to be due to a lack of understanding. For example, participants argued that people who were not observing social distancing lacked knowledge over how they could help spread the disease even if they themselves were not exhibiting symptoms. Third, non-adherence was seen to be due to a lack of enforcement. Many participants were critical that police were choosing to enforce social distancing restrictions or were not able to (due for example to the ambiguity of terms such as “essential” as discussed in the previous section). Others discussed how, despite their best efforts, supermarkets appeared to struggle to implement social distancing.

**Box 3: Current adherence and non-adherence of self and others**

*High levels of support for, and adherence to social distancing and isolation*

“Staying at home is actually helping to save lives” (Participant 20, female, age 21, South Wales)

“We have been in lockdown for 14 days, and because of my 87-year-old grandmother who has health problems, it [going out] is just not worth it.” (Participant 17, male, age 22, South Wales)

*Non-adherence due to lack of social conscience*

“I’m worried that people are going to take advantage of the nice weather and ruin it for people … Its insane because they have shut the park, but you get some inconsiderate people like a group of lads playing football or people taking over the paths.” (Participant 27, female, age 46, London)

“The canal path we walk along is not 2 metres wide, but you can just about get around it if you go on the verge and they go on the verge, and most people do but not everybody does … I don’t say anything because … with all the publicity that’s out, if you are still choosing to do that, then me telling you not to do it is not going to make a difference, it’s frustrating” (Participant 26, female, Manchester)

*Non-adherence due to lack of understanding*

The vast majority of people are taking it seriously and suffering to a certain extent, but there is a minority who don’t necessarily understand it applies to them also. I know of people who have gone to parks or gone for a picnic, because they think ‘well we don’t know anyone who has any symptoms, and we’ve not got anything, so we can go about it in the same way’.” (Participant 19, female, age 21, South Wales).

*Non-adherence due to lack of enforcement*

“They say that you are not allowed to go out for non-emergency reasons, which I don’t think a lot of people are observing. People are just going out whenever they want. Those guidelines are in place by Boris [Johnson; UK Prime Minister] but no-one is really enforcing that. You see police on the street, but they are not really doing anything.” (Participant 2, male, age 31, Manchester)

“The supermarket they are not implementing, what’s the point in having the two-metre thing outside when you can’t do that inside. … I went to the supermarket and people respect it outside, but as soon as you go inside there is [sic] people, they don’t care, they just come right up to you and try to reach over you.” (Participant 8, female, age 40, London).

### Uncertainty, social-reintegration and the future

According to participants, “the biggest problem we’ve got is we don’t know when it’s going to end” and the sense of “powerlessness” this had fostered (Participant 25, male, age 58,). Despite their high level of current adherence, participants acknowledged there was a limit as to how long they and others could adhere, at least without experiencing more severe social and psychological suffering. Some participants felt that they would rather be told a specific time frame, even if it was far in the future. Others feared that whilst they and others could “get through” this initial phase of lockdown, going “in and out” of periods of lockdown (a scenario some knew was possible due to the potential for COVID-19 to re-emerge in a second wave) meant that “people will really struggle mentally” (Participant 19, female, age 21, South Wales). Some felt as though they could only take things “day-by-day” because anticipating social distancing and isolation over a period of time was “too overwhelming” (Participant 22, male, age 51, North-West England).

Looking to the future, participants were divided as to how they felt they, and others, would act when social distancing and isolation measures were either relaxed or removed. Some felt that they and others would “go back to living my life completely as normal” (Participant 24, male, Manchester) as soon as possible. These participants spoke of “being desperate to go out and go to restaurants or travel a lot” (Participant 5, male, age 26, London) and generally not taking a graded approach to social reintegration. They argued that if they were “told its ok” to socially reintegrate, then this was enough for them to “not feel too anxious about going out with friends in the future” (Participant 24, male, age 40, North-West England). Others felt that it would take them a longer to return to pre-pandemic social behaviours, and for example felt that they would continue to have “anxiety around health” (Participant 23, female, age 38, North-West England), would be “cautious” about a “transition period where I stay in a bit more” (Participant 2, male, age 31, Manchester) and that people in general might remain “socially distant” from one another (Participant 8, female, age 40, London).

Others argued that how they would act would likely depend on the circumstances under which social distancing and isolation measures were being relaxed or removed.

Specifically, this was tied to their perception of whether COVID-19 still posed a risk to them or to society in general. They argued that, if a vaccine was available, then they would be happy to return to their pre-pandemic activity.

**Box 4: Uncertainty, social-reintegration and the future**

*The challenge of future uncertainty*

“I would rather they [the government] said tonight, ‘you’re gonna be stuck in your houses until September, than say, we will review in three weeks, and then say, we will review in three week, and keep doing that, I’d rather they set a date way in advance in the future because then you can get your head around it” (Participant 17, male, age 22, South Wales)

“I’ve heard on the grapevine and online sources that we are in this lockdown for a few weeks or so, and then after 12 weeks or so we kind of get released and because we are not all immune necessarily it all comes back in a wave and then we have this constant thing of being locked down and then coming out and going back in again, … and so I think it will go downhill, that’s when people will struggle mentally because they’ve had that taste of freedom, and you don’t know how long it’s all going to finish.” (Participant 19, female, age 21, South Wales)

*Perceptions of future behaviour*

“I’m literally thinking day-by-day, because if one was to consider three months of this, and we are only two weeks in, it’s just too much, it’s too overwhelming” (Participant 22, male, age 51, North-West England)

“People are not going to stay like this for another 6 months. It’s for a good reason I know, but it’s like a prison, we know what people are suffering mentally and emotionally, we don’t know what people are going through behind their door … When all this comes to an end we don’t know how life is going to be. Is everybody going to be socially distant? It’s scary.” (Participant 8, female, age 40, London)

### Alternative accounts and positive perceptions resulting from social distancing and isolation

Although the findings discussed above represent the most common views exhibited by participants, negative case analysis did reveal some alternative accounts. For example, some participants argued that social distancing and isolation “hadn’t been hard” (Participant 17, male, age 22, South Wales). However, these participants were all university students, and acknowledged that part of the reason it hadn’t been as difficult for them was there had been no loss of income and, less loss of routine for them.

A small number of participants argued that they were able to draw positives from the social distancing and isolation due to COVID-19. For example, some described how household quarantine had meant they could have “more time with their children” (Participant 14, male, age 38, North-West England) or had brought family units together (Participant 22, male, age 51, North-West England). However, those participants who explicitly discussed the positives to be drawn from social distancing and isolation were all from higher socio-economic backgrounds, and tended to live in more rural or less densely populated areas of the UK.

## DISCUSSION

Our findings suggest that a large proportion of the UK public may be suffering from feelings of depression, anxiety, and loss as a result of COVID-19 social distancing and isolation. Some already feel in need of professional mental health support. The social and psychological impacts identified through this study centred around the various losses that people are experiencing. Practical social and economic losses - the loss of (in-person) social interaction, loss of income and loss of structure and routine – led to psychological and emotional losses – the loss of motivation, loss of meaning, and loss of self-worth. Findings also suggest that participants generally found information on social distancing to be ambiguous. However, there were differing views as to whether the government was at fault (insufficiently clear communication) or that the terms themselves are ambiguous and a small minority of the UK public were taking advantage of the ambiguity. It was recognized that this ambiguity may have been designed to permit greater social freedom than the more extreme social distancing and isolation measures implemented in other countries. Additionally, there was universally high adherence to social distancing and isolation guidelines reported across the study sample, yet most participants had observed or heard of non-adherence in others. Participants were highly critical of such instances of non-adherence, citing lack of social conscience, lack of understanding and lack of enforcement as likely causes. Perhaps the greatest concern for participants was the uncertainty they faced over the duration of the social distancing and isolation measures, as well as their ability to cope longer-term. There was also uncertainty as to how they and others would act, with some fearful of lingering inhibitions and anxiety over social contact and health, and others eager to return to normal levels of social activity.

### Relevance to existing literature

Our findings on COVID-19 social distancing and isolation support some of the findings from existing systematic reviews on previous pandemics related to influenza and SARS.[6-7] For instance, we found that frustration or anxiety over loss of social interaction or loss of income, inadequate or ambiguous information, and fears over the duration of social distancing and isolation measures were all major themes.[6-8]. However, contrary to previous research which suggests that adherence with pandemic NPIs is lower in instances where people have low trust in government and where people perceive themselves at relatively low risk from the disease,[7] our participants were highly adherent to social distancing and isolation measures, despite many lacking trust in government and perceiving themselves at low risk. In fact, stigma was more likely to be attributed to those who were failing to socially distance and isolate. Of course, as noted above the scale and severity of the pandemic and subsequent measures are unprecedented. As such, although there is existing research on its likely effects,[6] and although quantitative research is starting to emerge,[11] there is to, our knowledge, no published qualitative evidence on public perceptions and experiences of the psychological and social public experiences of COVID-19 related social distancing and isolation, and its relation to adherence – a gap this study addresses.

### Limitations

One limitation of this study is that it is not possible to rule out that the high degree of adherence and social conscience that participants expressed was not at least partly affected by social desirability bias, which can often be encountered in focus group studies.[19] However, conducting focus groups online has been found to reduce social desirability bias (although it is worth noting that this is more so where asynchronous or text-only communication is used, and not video-conferencing as in our study).[12,20]

Another limitation of this study is that it did not recruit participants who are deemed at particularly high risk from COVID-19-related complications, for example, individuals aged 70 and over and those living with certain chronic health conditions.[21] Because these individuals are likely to have been significantly affected by social distancing and isolation policy (being required to self-isolate for 12 weeks), their views will be important. It is also worth noting that our recruitment material did encourage those at high risk to apply, though we received no applications from those over-70. This may be partly due to the fact that those over-70 are a hard-to-reach group online, because they are significantly less likely to use social media or be heavy internet users,[22] which, due to the lack of online social support and interaction, might mean they are at particularly high risk of some of the negative social and psychological impacts discussed in this paper. Future research will explore at-risk groups’ experiences in depth. Future papers will also explore further the similarities and differences in views and experiences in the perceptions of experiences of participants living in different parts of the UK (e.g. London compared to less densely populated areas), a theme only briefly discussed here due to limitations of scope.

### Implications for policy and practice

This study suggests that the social distancing and isolation associated with COVID-19 policy is having substantial negative impacts on the mental health and wellbeing of the UK public within a short time of policy implementation. The prevalence of COVID-19-related depression and anxiety, and the extent to which it will last beyond the removal or relaxation of social distancing and isolation policies remains to be seen. Our ongoing research will explore these social and psychological impacts longitudinally. Policymakers and the public health community must discuss measures to respond to the likely wave of mental ill-health which is expected to follow, and which is tentatively suggested by our early qualitative evidence. The theme of loss and addressing public concerns around physical and emotional losses (e.g. meaning and self-worth), may inform current and future therapeutic interventions. Loss of meaning and self-worth may be due in part to loss of control, and increasing a sense of control for the public should be considered in future policy, intervention, and programming.[23] Additionally, findings suggest that a rapid response is necessary in terms of public health programming to mitigate these mental health impacts. Waiting until restrictions and isolation measures are relaxed or removed to provide support services could potentially have devastating impacts. Government and the public health authorities should look at ways of extending mental health outreach services, especially remotely.[24] Timely attention is needed for those who are predisposed to depression and anxiety, those who may be suicidal, and those experiencing significant social, economic and personal loss.

Our study also suggests that although the COVID-19 pandemic has had significant ramifications for many UK residents from diverse backgrounds, it has disproportionately negatively affected those in low-paid or precarious employment. Future research and policy should therefore seek to develop measures that specifically seek to remediate the social, economic and psychological harms related to COVID-19 as experienced by those from disadvantaged backgrounds. Looking ahead to later stages in the current pandemic, or to the development of pandemic preparedness programmes for the future, a couple of lessons can be distilled, which warrant urgent attention. Firstly, initial high levels of support for, and adherence to, social distancing and isolation measures are likely to wane over time, particularly where end dates are and remain uncertain. Secondly, in planning the ‘exit strategy’ for the UK lockdown, and its possible impact on future resurgences of COVID-19 infection, policymakers and public health authorities need to account for the fact that, although some individuals will voluntarily or habitually continue to socially distance (graded social reintegration) others will seek immediately to re-integrate fully beyond what they are permitted to.

## Data Availability

Ethical restrictions related to participant confidentiality prohibit the authors from making the data set publicly available. During the consent process, participants were explicitly guaranteed that the data would only be seen my members of the study team. For any discussions about the data set please contact the corresponding author, Simon Williams.

## DECLARATIONS

### Competing interest statement

All authors have completed the Unified Competing Interest form (available on request from the corresponding author) and declare: Armitage is supported by NIHR Manchester Biomedical Research Centre and NIHR Greater Manchester Patient Safety Translational Research Centre. Tampe is an independent consultant and currently consults for the World Health Organization. The authors have no other relationships or activities that could appear to have influenced the submitted work.

### Transparency declaration

The lead author (the manuscript’s guarantor) affirms that the manuscript is an honest, accurate, and transparent account of the study being reported; that no important aspects of the study have been omitted; and that any discrepancies from the study as planned (and, if relevant, registered) have been explained.

### Authors’ contributions

All authors contributed to the planning of the study. The analysis was conducted by SW and KD. The initial draft of the article was written by SW. All authors revised the manuscript and approved the final version for publication. SW is the guarantor of the article.

### Funding statement

This research was supported by the Manchester Centre for Health Psychology based at the University of Manchester (£2000). Armitage’s contribution was additionally supported by the NIHR Manchester Biomedical Research Centre and the NIHR Greater Manchester Patient Safety Translational Research Centre. The funders played no role in the conduct of the study.

### Ethics statement

Ethical approval was received by Swansea University’s Research Ethics <u>Committee.</u>

